# BACTERIAL ONCOTRAITS BUT NOT BIOFILMS ARE ASSOCIATED WITH DYSPLASIA IN ULCERATIVE COLITIS

**DOI:** 10.1101/2022.09.09.22279675

**Authors:** Carlijn E. Bruggeling, Maarten te Groen, Daniel R. Garza, Famke van Heeckeren tot Overlaer, Joyce P.M. Krekels, Basma-Chick Sulaiman, Davy Karel, Athreyu Rulof, Anne R. Schaaphok, Daniel L.A.H. Hornikx, Iris D. Nagtegaal, Bas E. Dutilh, Frank Hoentjen, Annemarie Boleij

**Author notes:** **Corresponding author:** Dr. Annemarie Boleij, Radboudumc, Department of Pathology (hp824), Geert grooteplein Zuid 10, 6525 GA Nijmegen, The Netherlands. Shared authorship (Maarten te Groen, Daniel R. Garza). **Author contributions:** study design (AB, FH), patient recruitment and sample processing (CEB, JK, FH), clinical data acquisition and analyses (AB, CEB, MtG, JK, IDN, ARS), metagenomics (CEB, DRG, BED), Biofilm assessment (CEB, FHO, JK, AR, AB), oncotrait analyses (BS, DK, AB), Mucus and ki67 assessment (CEB, AB, DLAH), pathology (IDN), supervision (AB, BED, FH), article writing (all authors). **Data Availability:** Raw sequencing data with human reads will not be publicly available because of General Data Protection Regulation (GDPR). Processed sequencing data are available upon request. Patient and research data are anonymized and openly available in supplementary data.

## Abstract

Biofilms are polymeric matrices containing bacteria that can express oncotraits and are frequently present in ulcerative colitis (UC). Oncotraits can impact colon epithelial cells directly and may increase dysplasia risk. This study aimed to determine (1) the association of oncotraits and longitudinal biofilm presence with dysplasia risk in UC, and (2) the relation of bacterial composition with biofilms and dysplasia risk.

In this prospective cohort study, feces and left- and right-sided colonic biopsies were collected from 80 UC patients and 35 controls. Oncotraits (FadA of *Fusobacterium*, BFT of *Bacteroides fragilis*, Colibactin (ClbB) and Intimin (Eae) of *Escherichia coli*) in fecal DNA were assessed with multiplex qPCR. Biopsies were analyzed for biofilms (n=873) with 16S rRNA fluorescent *in situ* hybridization and shotgun metagenomic sequencing (n=265), and ki67-immunohistochemistry for cell proliferation. Associations were determined with a regression (mixed) model.

ClbB significantly associated with dysplasia in UC (aOR 7.16, (95%CI 1.75-29.28, p<0.01)), while FadA was inversely associated (aOR 0.23, (95%CI 0.06-0.83, p=0.03)). Patients with UC had a significantly lower Shannon diversity compared to controls (p=0.0009), as well as patients with a biofilm (p=0.015) independent of disease status. The order *Fusobacteriales* was significantly correlated with a decreased dysplasia risk only in right-sided colonic biopsies (p<0.01). Longitudinal biofilms were not significantly associated with dysplasia (aOR 1.45 (95% CI0.63-3.40, p=0.38)), however, biofilm-positive biopsies showed increased epithelial hypertrophy (p=0.025).

Colibactin and FadA impact dysplasia risk in UC, in contrast to biofilms. These oncotraits are valuable targets for future risk classification and intervention studies.

**What is already known on this topic:** Bacterial biofilms sometimes contain bacteria with oncogenic traits (oncotraits) and have been associated with colon carcinogenesis in mice and humans. It is yet unknown whether biofilms and oncotraits are involved in early carcinogenesis and could be used as a risk factor for dysplasia in ulcerative colitis patients.

**What this study adds:** Bacterial biofilms associated with lower bacterial diversity and epithelial cell hypertrophy, but did not predict dysplasia. Moreover, in agreement to piling evidence suggesting a role of colibactin in human colorectal cancer, we provide the missing clinical evidence that this oncotrait actually associates with risk for (early) carcinogenesis in human patients. Additionally, dysplasia in UC patients was predicted by absence of Fusobacterium adhesin.

**How this study might affect research, practice or policy:** This prospective cohort study indicates a putative role of bacterial oncotraits in early carcinogenesis, suggesting them as promising targets for future risk classification and intervention studies in ulcerative colitis patients.

**Lay summary:** Patients with ulcerative colitis have an increased risk for colorectal cancer. This study found that bacterial factors in fecal material can predict the development of cancer precursors in these patients.

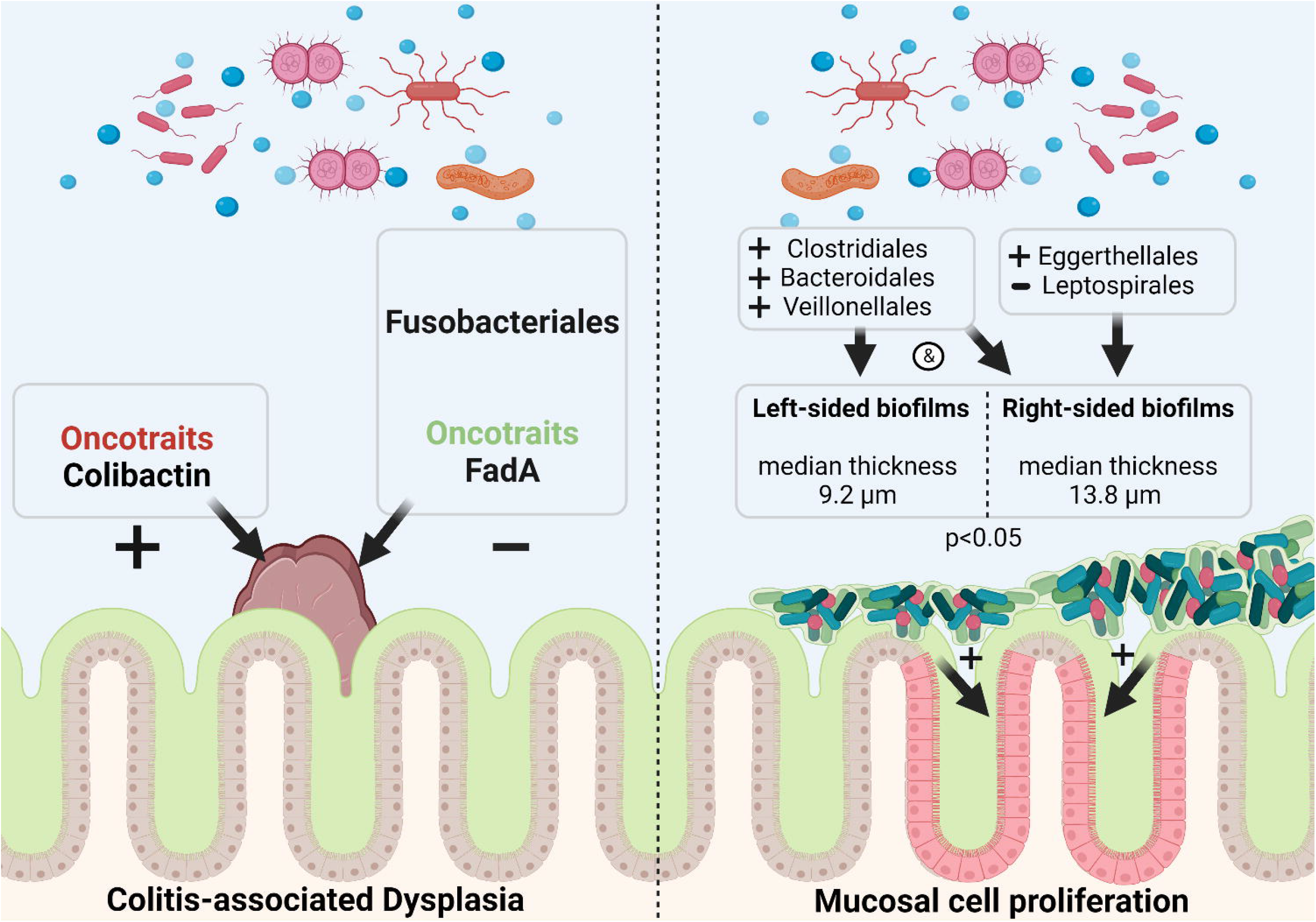

## Introduction

Ulcerative colitis (UC) is characterized by chronic inflammation of the colonic mucosa and is an increasing burden worldwide.[1] In UC chronic inflammation may lead to the development of mucosal dysplasia that can progress to colorectal cancer (CRC).[2,3] Detection of dysplasia and risk stratification are dependent on invasive techniques including surveillance colonoscopies every 1-5 years. Cancer prevention in UC is limited by our knowledge on accurate predictors of dysplasia and human errors in dysplasia detection, with 30-55% of CRC diagnosed as interval lesions.[4-6] Current non-invasive tests for CRC detection are based on fecal occult blood or hemoglobin detection, which is unreliable in UC because of high false-positive rates due to inflammation related bleeding. Investigation of novel biomarkers might provide new leads for less-invasive risk stratification. The colonic microbiota and its interaction with the (impaired) mucosal barrier may play a role in colitis associated cancer development and could potentially provide novel biomarkers for dysplasia in UC.

The inner mucosal layer of UC patients is frequently covered with bacteria in an adherent polymeric matrix, so-called biofilm.[7,8] Cross-sectional studies have shown endoscopic visibility of biofilms in 34% of UC patients[7], and microscopic detection in 70% of UC patients.[8] Bacteria frequently detected in mucosal biofilms in UC include *Bacteroides fragilis, Escherichia coli Klebsiella sp*., *Fusobacterium peridonticum*and *Ruminococcus gnavus*.*[7,8]* Biofilms provide a microenvironment for bacteria to thrive on the mucosal surface. Such bacteria may be pathogenic by carrying or producing pro-oncogenic bacterial products, so called oncotraits, as recently discovered in familial adenomatous polyposis.[9] Dejea et al., demonstrated that two oncotraits, colibactin from *Escherichia coli* and *Bacteroides fragilis* toxin (BFT), co-occur in biofilms and combined cause tumor formation in mice. Besides BFT and Colibactin, intimin (Eae) of *E. coli* and *Fusobacterium* adhesin protein A (FadA) have also been linked to CRC development.[10-14] These oncotraits are detectable in feces and bear potential as biomarker for an increased dysplasia risk or presence of dysplasia.[15] However, the exact role of oncotraits and bacterial biofilms and their potential as biomarkers of carcinogenesis in UC is unknown.

In this study we analyzed longitudinal colonic biopsies and fecal samples from UC patients undergoing surveillance to determine (1) the association of oncotraits, and longitudinal biofilm presence with dysplasia risk in UC, (2) bacterial composition and its relation with biofilms, and dysplasia risk.

## Materials and Methods

### Study design

We performed a prospective cohort study to assess associations of oncotraits and biofilms with dysplasia, bacterial composition and mucosal cell proliferation. We employed an additional retrospective longitudinal arm for in-depth biofilm characterization of the included UC patients. For the additional longitudinal biofilm assessment, only high-quality data was included, that is, (1) patients with at least 3 colonoscopies including the study procedure and (2) colonoscopies with at least left- and right-sided biopsies. Longitudinal data of the UC patients was gathered up to the moment of the study colonoscopy.

### Study population

Patients were included at the Gastroenterology department (Radboud University Medical Centre, The Netherlands) in a consecutive manner from December 2016 to September 2018. Cases with UC were included if they had (1) disease duration longer than 8 years, left-sided colitis or extensive disease (Montreal score E2 or E3), and (2) before colonoscopic surveillance in clinical remission (Montreal score S0) for patients with UC. Exclusion criteria were antibiotics in the last 3 months before colonoscopy. Control group patients were included based on an indication for a colonoscopy for unexplained complaints of the intestine, such as changed bowel habits, unclarified abdominal pain, iron deficiency anemia or fecal blood loss. Exclusion criteria for this group were an IBD diagnosis, a history of CRC, colonic surgery, or antibiotics in the last 3 months before colonoscopy, abnormal findings during study colonoscopy (Figure 1). The study (NL55930.091.16) was approved by the Internal Revenue Board CMO-Arnhem Nijmegen (CMO 2016-2818) and the board of the Radboudumc. All participants gave written informed consent.

**Figure 1.**
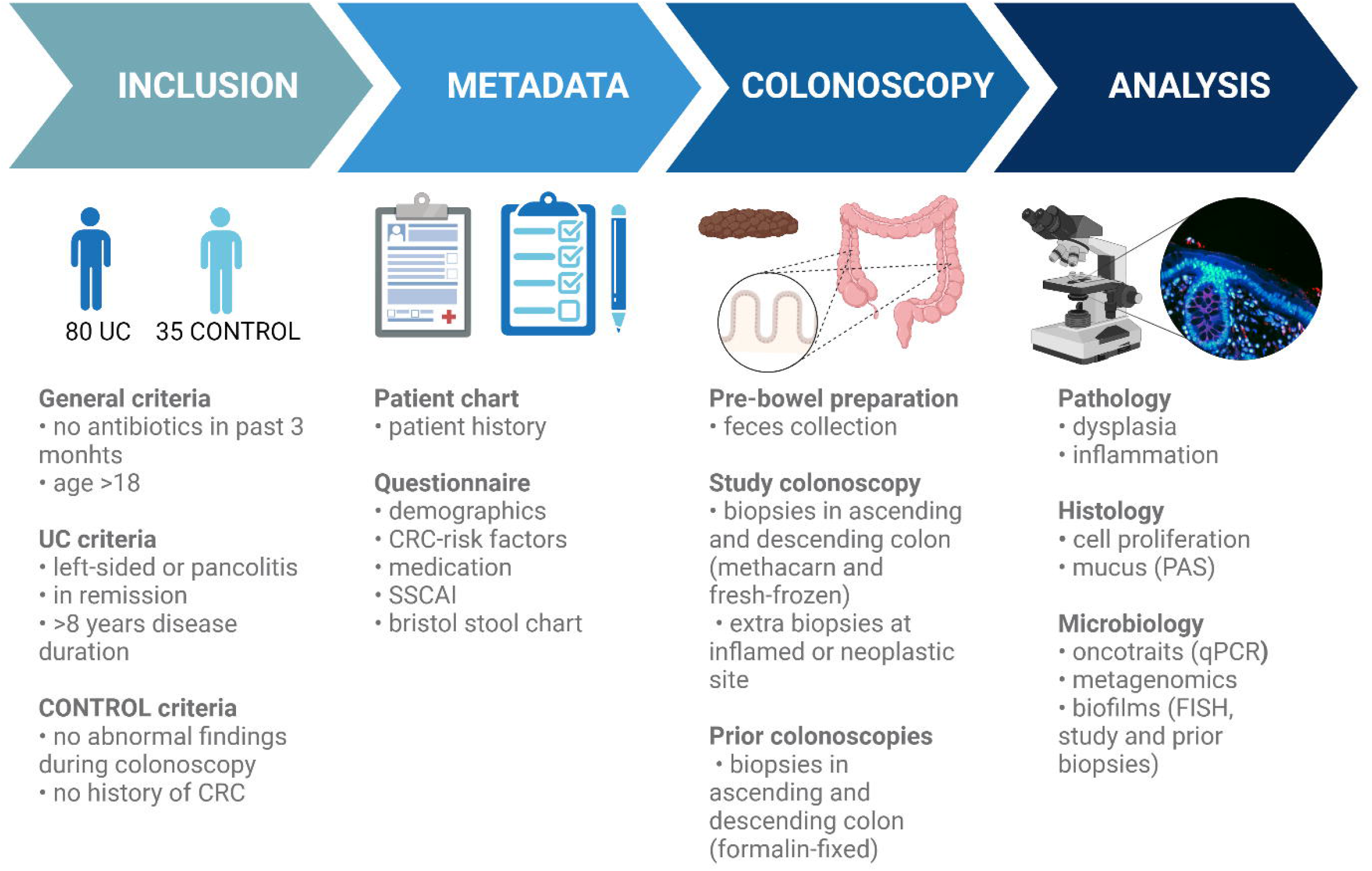
Study workflow from inclusion to collection of data and samples to analysis.

### Clinical parameters

Metadata collected included patient’s disease history, demographics, CRC-risk factors, (IBD-relatd) medication, SSCAI, Bristol stool chart, which were extracted from patient files, and questionnaire.

### Tissue and stool sample collection

Patients underwent a study colonoscopy between December 2016 and December 2018 in the Radboudumc Nijmegen. Patients were asked to bring a cooled home-collected fecal sample to the appointment, with the instruction of collecting it shortly before bowel cleansing (the morning or early afternoon the day before the colonoscopy). Fecal samples were homogenized, aliquoted and stored at -80°C. From all patients, biopsies were taken from the ascending and descending colon and, when present, at the site of a suspected dysplastic lesion. All biopsies were collected according to 3 different methods, 1) fixed in methacarn (60% methanol, 10% acetic acid and 30% chloroform), 2) formalin, and 3) snap-frozen. Snap-frozen biopsies were stored at -80°C until use. Endoscopic assessment of the severity of inflammation was performed for each biopsy location using the endoscopic Mayo score. In addition, retrospective data from prior colonoscopies was collected through patient files.

### Pathology

Prospectively collected biopsies from UC and control patients fixed in formalin and methacarn were processed for paraffin embedding at the Radboudumc pathology department and pathology diagnosis were retrieved form patient files. Retrospective FFPE tissue biopsies from UC patients were gathered from the Radboudumc archives up to the first registered screening colonoscopy, resulting in 402 colonoscopies of which we analyzed 873 biopsies from the 1240 available biopsies. Pathology data, such as inflammation score (mild, moderate, severe), and dysplasia grade (low grade, high grade) were extracted from pseudonymized pathology reports. Methods for tissue processing for Periodic Acid Schiff (PAS), IHC, and FISH, and corresponding analysis are provided in the Supplementary data 1.

### Bacterial DNA isolation

Bacterial DNA was isolated using a previously published, optimized method.[16] Fecal (200mg) and tissue DNA from snap frozen biopsies was isolated with a modified HMP protocol optimized for bacterial DNA analysis[17], which included mutanolysin (SAE0092, Sigma Aldrich) and bead-beating with the DNAeasy powerlyzer Powersoil kit (12855-50, Qiagen) and was extended with additional proteinase K (19133, Qiagen), Saponin (47036-50G, Sigma-Aldrich), DNAse mediated enrichment (AM2239, Qiagen) for tissue specimens. Methods for metagenomics and corresponding bioinformatic analysis is provided in Supplementary data 1.

### Biofilm scoring

Mucosa associated bacteria were visualized with a universal 16S rRNA probe (EUB338 labeled with Cy3/Cy3) and fluorescence microscopy (Leica DMRA) (Supplementary data 1). The tissue was scored based on bacterial abundance and biofilm formation. Tissue was scored in 4 tiers: no bacteria (0), low (1), moderate (2) or high (3) bacterial abundance on the mucosa. Only tissues which were assessed moderate (2) or high (3) were scored for biofilms and a representative picture was taken. Biofilms were defined as a continuous plaque of bacteria covering at least 100 μm of the epithelial surface and were measured with Fiji (version 1.51n). All biopsies from the study colonoscopy were scored by two independent observers and a third observer was consulted in case of disagreement to reach consensus. For the longitudinal retrospective biopsies, a random selection of 174 of the 873 biopsies were scored by a second observer. The agreement was 86.8% resulting in a kappa of 0.63 with substantial agreement (95% CI 0.48-0.77); Supplementary Table 1).

### Oncotrait prevalence

To detect the bacterial oncotraits BFT from *Bacteroides fragilis*, colibactin (ClbB) and intimin (Eae) of *Escherichia coli*, and FadA of *Fusobacterium sp*., a multiplex PCR was performed on fecal DNA (Supplementary data 1).

### Metagenomics analysis

Metagenomic sequencing and analysis were performed as previously described (See Supplementary data 1 and Supplementary Table 2).[16]

### Statistics

The number of patients needed for the study was based on previous observations considering the rate of bacterial biofilms in UC patients (70%)[8] versus controls (15%)[18], and the rate of BFT in controls (51.3%) and CRC patients (88.5%).[19] To find a statistical difference in proportions with Chi-square statistics between patients with dysplasia and without 19 patients per group are needed. As only 70% presents with a biofilm, 27 patients per group are minimally needed. Based on these calculations and the possibility to at least correct for 1 confounder in binary logistic regression we aimed to include 40 control patients, 40 UC patients with (history of) dysplasia and 40 UC patients without dysplasia. Statistical tests were performed using Graphpad Prism v9.0.0, GraphPad Software LLC, USA) and IBM SPSS Statistics v25 (Armonk, NY, USA). Descriptive statistics were assessed, and Chi-square tests were performed and displayed with 95% confidence intervals. Correlations were assessed with Pearson’s or Spearman’s tests. A binary logistic regression model was used to assess (1) univariable associations with dysplasia, (2) adjust for confounders and (3) independency of found associations.

A regression framework was made using Daggity (Supplementary Figure 1).[20] Factors were selected for the multivariable model based on expert opinion and clinical relevance. In addition, to adjust for repeated measures a binary logistic mixed model was used with subjects as random effects to assess longitudinal biofilm data. Two-sided p-values <0.05 were considered statistically significant. For the cross-sectional data, we employed dysplasia at the study colonoscopy or in the prior five years as a composite endpoint. The following definitions were used for comparison of the longitudinal data: right-sided colon included coecum and ascending colon, whereas left-sided colon included descending colon and sigmoid. Rectal biopsies were excluded because of sparse data. In biofilm characterization and metagenomics data in relation to dysplasia, high-risk UC patients were considered those with current or previous dysplasia, or concomitant primary sclerosing cholangitis (PSC). All other patients were considered low-risk UC.

## Results

### Patient demographics

#### Baseline characteristics

In total, 115 patients (n=80 UC patients and n=35 controls) were included after exclusion of 11 controls with abnormal colonoscopy findings (low grade dysplasia, inflammation, microscopic colitis, and spirochetosis). UC patients were more often male compared to controls (58.8% vs 31.4%, p<0.01), had a diagnosis of pancolitis (77.2%), PSC (16.3%) and a median disease duration of 21 years (IQR 12.5-29.5). Of these, 68.8% used amino-salicylates and 17.5% were on biological therapy. The median retrospective follow-up time was 7.5 years (IQR 4.5-11.0 years). Details of the study selection and baseline characteristics are displayed in Figure 1 and Table 1-2.

**Table 1.**
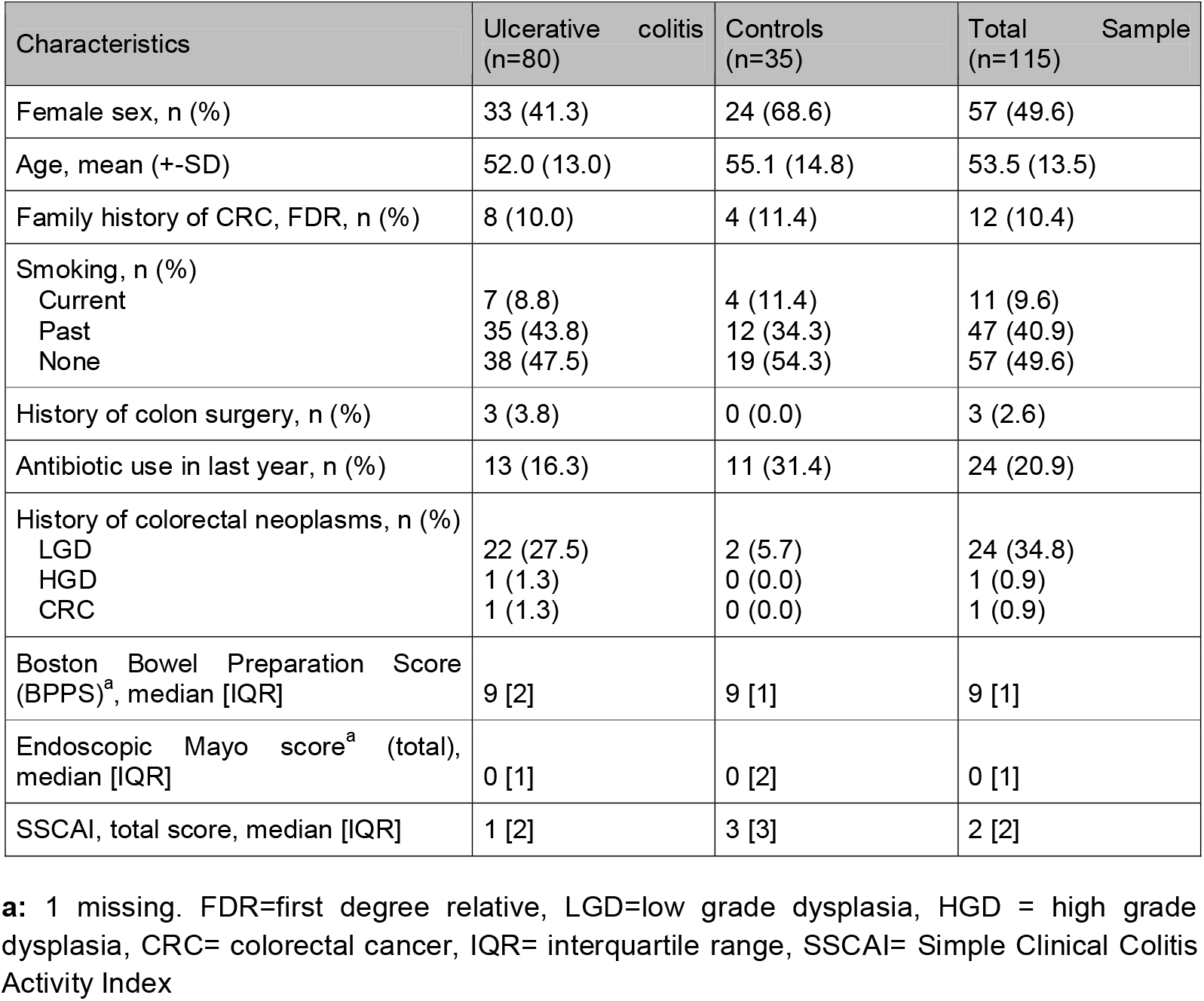
Baseline characteristics for ulcerative colitis patients and controls*.

**Table 2.** UC specific baseline characteristics

| UC specific baseline characteristics |  |
| --- | --- |
| Age at UC diagnosis, mean [±SD] | 31.2 [12.5] |
| PSC, n (%) | 13 (16.3) |
| History of post inflammatory polyps, n (%) | 19 (23.8) |
| <100 | 14 (66.6) |
| >100 | 5 (6.3) |
| Number of colonoscopies<sup>a</sup>, median [IQR] | 4 [4] |
| Follow-up after UC diagnosis until index colonoscopy, median years [IQR] | 21 [17] |
| Maximal endoscopic extent (Montreal)<sup>a</sup>, n (%) |  |
| E2 | 18 (22.8) |
| E3 | 51 (77.2) |
| Maximal histological extent (Montreal)<sup>b</sup>, n (%) |  |
| E2 | 19 (24.4) |
| E3 | 49 (62.8) |
| Unknown | 1 (1.3) |
| Maximal endoscopic inflammation severity, n (%) |  |
| Mild | 18 (22.5) |
| Moderate | 17 (21.3) |
| Severe | 25 (31.3) |
| Unknown | 20 (25.0) |
| IBD medication<sup>c</sup>, n (%) |  |
| Aminosalicylates | 55 (68.8) |
| Immunomodulator | 17 (23.0) |
| Biological | 13 (17.5) |
| Prednisone | 8 (13.5) |

**a:** 1 missing. **b:** 2 missing. **c:** 6 missing. PSC= primary sclerosing cholangitis, SD= standard deviation, IQR=interquartile range, IBD= inflammatory bowel disease.

#### Dysplasia

Dysplastic lesions identified in UC patients during the study colonoscopy were all classified as tubular adenomas with low grade dysplasia and localized in the coecum (n=1), ascending (n=2), transversum (n=4), descending (n=2), sigmoid (n=1) and rectum (n=2) (58.3% right colon, 25% left colon and 16.6% rectum) (Supplementary Table 3 and 4). Of the 11 patients with dysplastic lesions during the study colonoscopy, 8 patients (72.7%) had a history of 1 or more dysplastic lesions. In addition, 9 patients had dysplasia in the five years prior, but not during the study colonoscopy. Disease duration (OR 1.06 (95%CI: 1.01-1.11)), and pseudopolyp presence (OR 1.80 (95%CI: 1.00-3.23)) were significantly associated with dysplasia development (p<0.05). Family history of CRC showed a non-significant increased odds (OR 3.50 (95%CI: 0.79-15.60), p=0.10). All univariable associations with dysplasia are displayed in Table 3.

**Table 3.** Univariable regression analysis for associations with dysplasia at the study colonoscopy or in the prior five years.

| Risk factor | OR (95% CI) | p-value |
| --- | --- | --- |
| Age (per year) | 1.02 (0.98-1.06) | 0.35 |
| Smoking (history or present) | 1.41 (0.61-3.22) | 0.42 |
| Medication |  |  |
| 5-ASA | 2.14 (0.63-7.35) | 0.23 |
| Thiopurine + MTX | 0.58 (0.17-1.99) | 0.39 |
| Anti-TNF | 0.56 (0.11-2.80) | 0.48 |
| NSAIDs | 1.56 (0.26-9.21) | 0.63 |
| Corticosteroids | 0.45 (0.05-3.91) | 0.47 |
| PPI | 1.49 (0.45-4.96) | 0.52 |
| Opioids | 3.50 (0.21-59.13) | 0.39 |
| Bile acid binders | 0.38 (0.04-3.30) | 0.38 |
| Biofilm presence |  |  |
| Any | 1.59 (0.50-5.10) | 0.43 |
| Left sided | 0.88 (0.29-2.70) | 0.82 |
| Right sided | 1.32 (0.47-3.71) | 0.60 |
| Extensive disease | 0.80 (0.27-2.33) | 0.68 |
| Familial history of CRC | 3.50 (0.79-15.60) | 0.10 |
| Pseudopolyp presence | 1.80 (1.00-3.23) | **0.049** |
| PSC | 0.50 (0.10-2.45) | 0.39 |
| Disease duration (cont., for each year) | 1.06 (1.01-1.11) | **0.03** |
| Oncotraits |  |  |
| Eae | 2.59 (0.73-9.2) | 0.14 |
| ClbB | 4.77 (1.36-16.71) | **0.02** |
| BFT | 1.06 (0.32-3.51) | 0.93 |
| FadA | 0.23 (0.06-0.84) | **0.03** |

OR: odds ratio, CI: confidence interval.

### Oncotraits

Oncotraits were assessed in fecal DNA of 59 UC patients (73.8%) and 28 controls (80%) who provided a fecal sample. One or more oncotraits were detected in 45 UC patients (76.3%) and 23 controls (82.1%). FadA was most frequently present in UC-patients (52.5%) and controls (50.0%; p=0.82), followed by BFT (39% UC vs 53.1% in controls; p=0.19) and ClbB (39.0% in UC vs 46.9% in controls; p=0.47) (Figure 2A and B). Thus, no difference was found in prevalence of oncotraits in UC compared to controls. While FadA and ClbB occur as single oncotrait in UC patients (33% and 26% respectively), Eae did not occur without ClbB, and BFT did not occur alone. In UC patients, FadA and BFT were the most frequently observed combination. In general, oncotraits rarely occurred alone in both populations (35.6% single vs 64.4% multiple oncotraits in UC, and 26.1% vs 73.9% in controls, p<0.05). In UC, ClbB was positively associated with dysplasia at the study colonoscopy or in the prior five years (OR 4.77 (95% CI 1.36-16.71), whereas FadA was negatively associated (OR 0.23 (95% CI 0.06-0.84; Table 3). After adjusting for disease duration, an aOR of 7.97 (95% CI 1.77-35.91) was observed for ClbB and aOR 0.15 (95% CI 0.03-0.67, p=0.01) for FadA. BFT and Eae were not associated with dysplasia in UC. To assess the independency of found associations an explorative model including both FadA and ClbB was used. The association of ClbB with dysplasia remained significant (aOR 4.0 (1.09-14.70).

**Figure 2.**
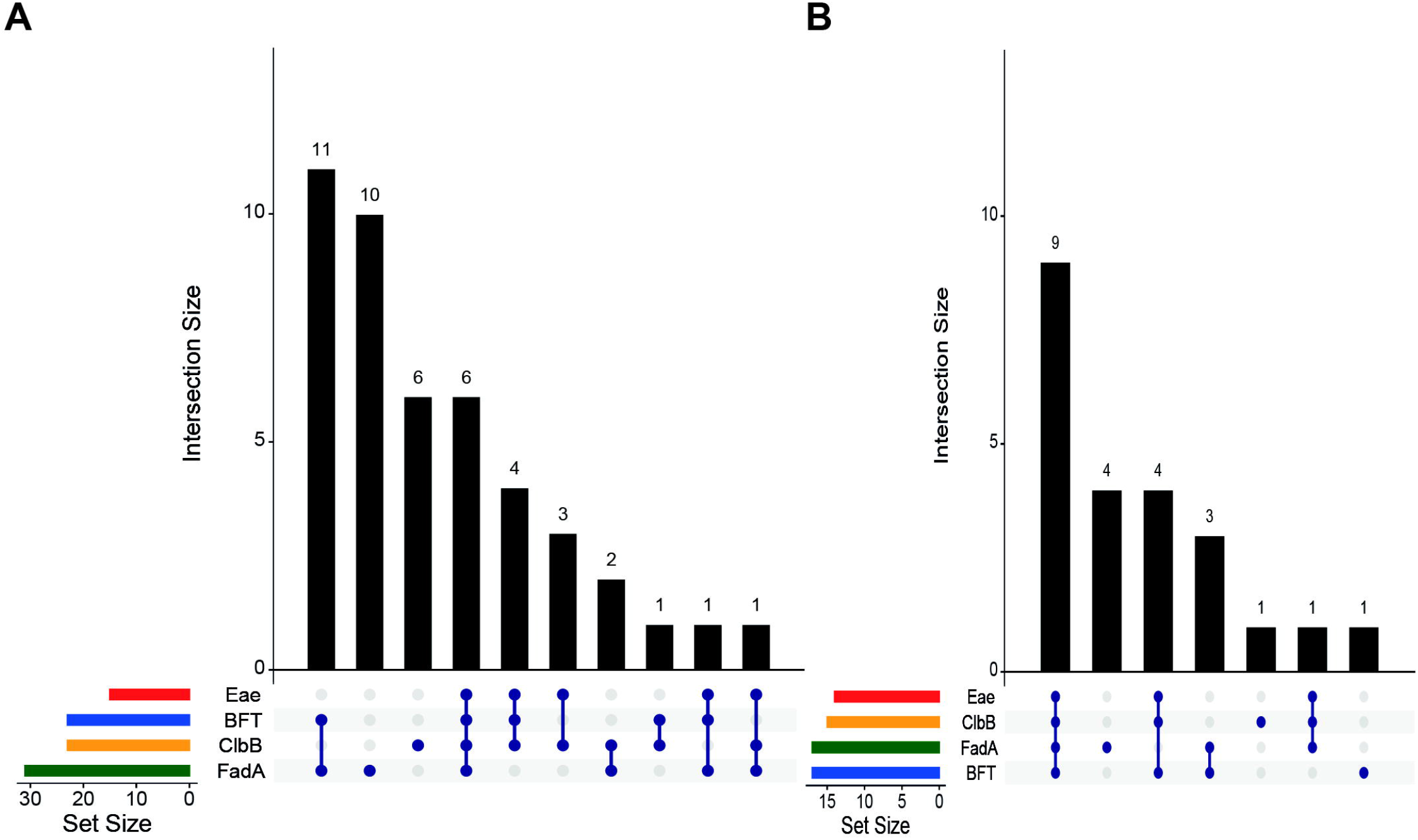
Frequency of oncotraits FadA (green), ClbB (yellow), BFT (blue) and Eae (red) in UC patients (A) and controls (B). Set size represents n patients detected with each oncotrait. Intersection size depicts the number of patients with the combinations of oncotraits (blue circles) on the y-axis.

### Biofilms

#### Biofilms at study colonoscopy

Biofilms (Figure 3A) were present in 50.0% of the controls at any location (left, and/or right colon; Figure 3B). In UC patients there was a slight increase in biofilm prevalence in subgroups of high-risk UC patients (72.2%): PSC (84.6%), dysplasia during endoscopy (70.0%), and any history of dysplasia (69.6%), compared to patients without (a history of) dysplasia or PSC (low-risk; 57.5%), but this was not significant (Figure 3B B; Supplementary Table 5). Biofilms occurred in 72.2% of patients with dysplasia at the study colonoscopy or in the prior five years (versus 62.1% in the remaining group, p=0.43). Biofilms occurred numerically more frequently in the ascending versus descending colon for both UC patients and controls (47 vs 36% in UC and 45 vs 32% in controls, p=0.08) (Supplementary Table 5).

**Figure 3.**
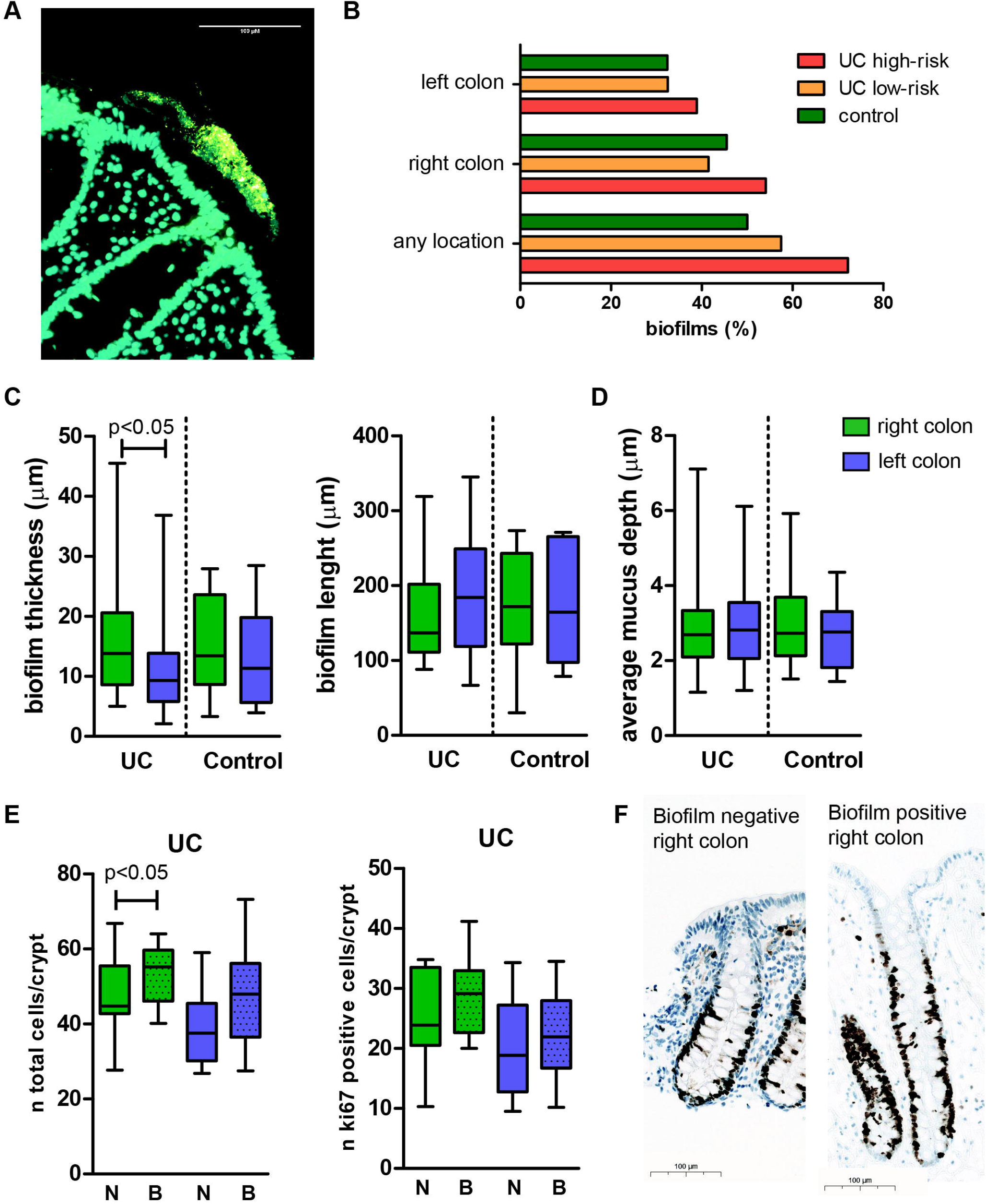
Biofilms and cell proliferation in ascending and descending colon biopsies. (A)Example of a bacterial biofilm spanning two colonic crypts in a UC patient. Cyan=nuclear staining (DAPI), yellow = Eubacteria staining (eub338) B) Prevalence of biofilms in ascending and descending colon biopsies at study colonoscopy in controls, low- and high-risk UC patients. C) Average biofilm thickness (µm) and biofilm length (µm) separated by left and right-sided colon biopsies in UC patients and controls. Right-sided colon = green, left-sided colon = blue. D) average mucus depth measured on PAS-stained right-(green) and left-sided (blue) colon biopsies. E) Number of cells per crypt and number of ki67-positive cells per crypt in biofilm positive (dotted boxplots) and biofilm negative (plain boxplots) in UC patients separated by right-(green) and left-sided colon (blue). N= no biofilm, B= biofilm F) Representative pictures of ki-67 staining (brown) in biofilm negative right colon and biofilm positive right colon of one UC-patient. Mann-Whitney U-test was performed for biofilm thickness and length, and mucus thickness; Independent students t-test was performed or number of cells per crypt and number of ki67 positive cells per crypt.

Biofilms in the right-sided colon were significantly thicker compared to left-sided biofilms (median 13.8 µm vs 9.2 µm, p=0.031) (Figure 3C), while there was no difference in average biofilm length. There were no significant differences in biofilm thickness, length and area between UC patients and controls (Supplementary Figure 2). No correlation was observed between average biofilm and mucus thickness in right (Spearman R 0.103) and left-sided colon biopsies (Spearman R 0.069) (Supplementary Figure 3). Biofilms were associated with epithelial hypertrophy in UC patients measured by the increased number of epithelial cells per crypt (median number of cells per crypt 42.5 vs 51.1, p=0.025), and a slight non-significant increase in Ki67-positive cells per crypt (22.0 vs 26.1, p=0.11) (Figure 3E and F).

#### Longitudinal biofilm characteristics in UC and associations with dysplasia

In 65 UC patients, 264 colonoscopies with left and right biopsies were performed. Left or right-sided biofilms were present at 141 colonoscopies (53.4%). Presence (at least on one occasion) of biofilms was observed in 59/65 (90.8%) of patients (Supplementary Figure 4A and B). The median duration of biofilm persistence in between biofilm-negative colonoscopies was 3 years (IQR 2-5 years) in patients with more than one biofilm-positive colonoscopy. Median number of colonoscopies in biofilm-positive intervals was 2.5 years(IQR 2-3.3 years). Biofilms were neither associated with endoscopically visible active inflammation in left and/or right-sided biopsies (p=0.43-0.92), nor with dysplasia (aOR after confounder correction for disease duration: 1.45 (95% CI 0.63-3.40, p=0.38).

#### Bacterial composition in UC patients and biofilms

Metagenomic shotgun analysis of left- and right-sided biopsies revealed that the bacterial composition in UC patients differed from those in the control population, characterized by a lower Shannon diversity index (4.80 vs 6.01 in controls; p=0.0009) (Figure 4A;Supplementary Table 6). Only in UC patients, right-sided biopsies displayed a significantly lower Shannon diversity as opposed to left-sided biopsies (Figure 4B), but was similar between UC patients with a low and high dysplasia risk. (Figure 4C).

**Figure 4.**
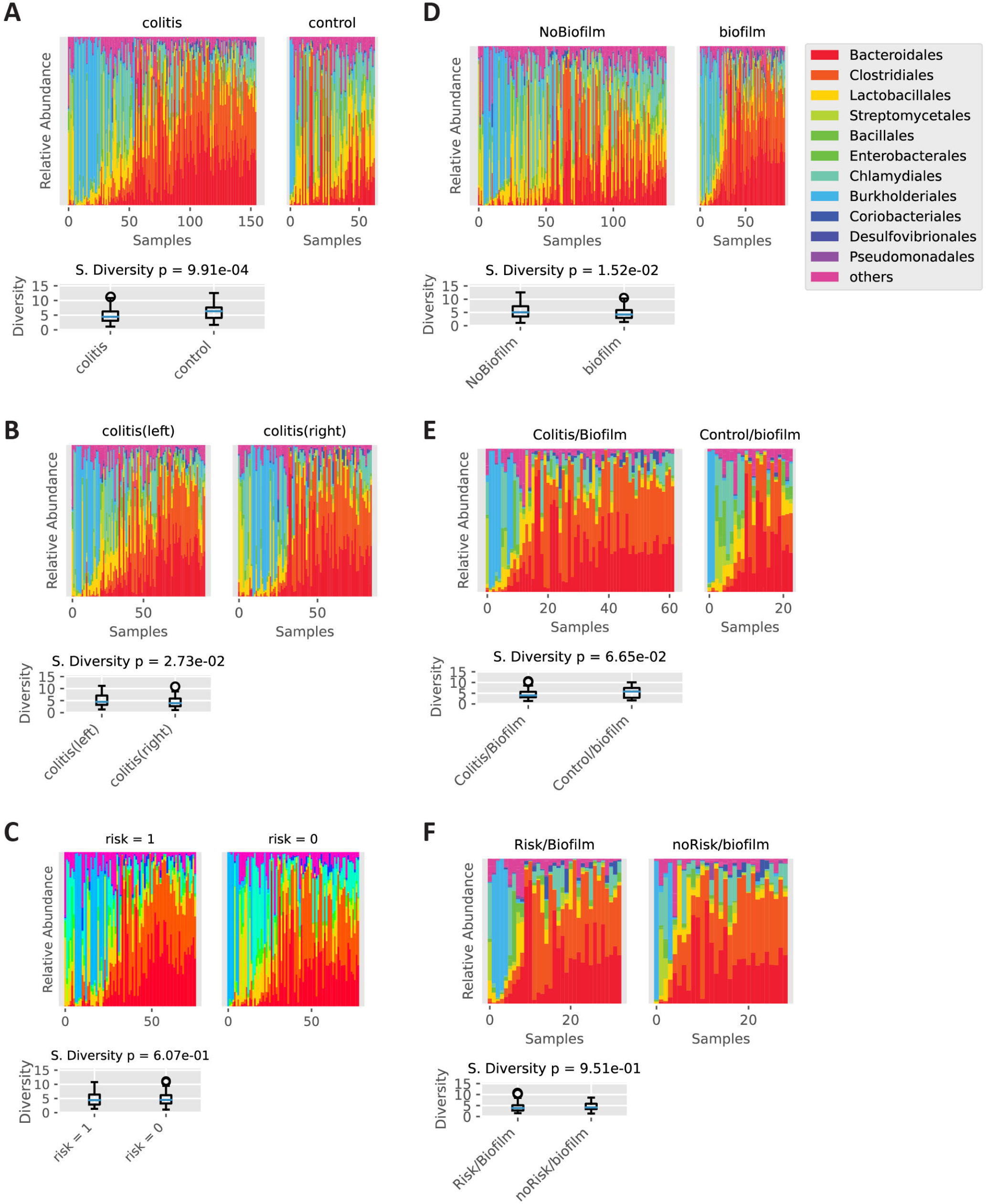
Metagenomic shotgun sequencing of healthy appearing colon ascendens (right) and descendens (left) in UC and control patients. Bacterial composition displayed at the order level with Shannon diversity (S. diversity). Samples were sorted by their correlation to the group’s mean composition: A) UC vs control patients, B) No biofilm vs biofilm of UC and control patients C) Left vs right colon of UC patients, D) Biofilms in UC vs control patients, E) High (1) vs low (0) risk UC patients in general, and F) Biofilms in high risk vs low risk UC patients.

Biopsies with biofilms displayed a lower Shannon diversity (p=0.0152; Figure 4D), but were not different between control and UC patients (Figure 4E), or between UC patients with low or high dysplasia risk (Figure 4F). ANCOM analysis showed that biofilm presence could be predicted by *Clostridiales, Bacteroidales* and *Veillonellales* (adj-p=0.012) in metagenomes of controls and *Selenomonadales* and *Synergistales* in metagenomes of UC patients (adj-p=0.011). More specifically, presence of *Eggerthellales* and absence of *Leptospirales* could predict biofilms in right-sided biopsies of high-risk UC patients (adj-p=0.023; Supplementary Table 7). Noteworthy is the observation that high risk for dysplasia in UC patients could be predicted by the absence of *Fusobacteriales* (p=0.00892; Supplementary figure 5), similar to the inverse relationship of FadA with high-risk UC patients.

## Discussion

Through this longitudinal study we provide unique evidence that supports a role of oncotrait colibactin (ClbB) in early carcinogenesis in UC patients. The commonly CRC-associated *Fusobacterium* along with its associated FadA was negatively associated with dysplasia in UC patients. Bacterial biofilms were studied longitudinally spanning 20 years in >800 biopsies of UC patients undergoing surveillance colonoscopies and were intermittently present in the majority of UC patients (90.8%). While longitudinal biofilms were not associated with dysplasia, their cross-sectional presence associated with a lower Shannon diversity and epithelial hypertrophy. Hence, our study does not support a general role for biofilms as risk factor for dysplasia in UC patients, but suggests that specific bacterial factors, such as presence of ClbB and absence of FadA could predict dysplasia risk in UC patients.

An altered microbiome composition and function has been associated with the development and progression of UC.[21] Currently, UC prediction algorithms based on microbial markers are rapidly developing[22], but these microbial markers have unknown roles in development and prediction of (early) carcinogenesis.[23] ClbB is located on the *pks+* island that encodes colibactin[24], which directly damages DNA and results in a mutational signature in human intestinal organoids that can be found in some CRC patients.[10] Thusfar, the direct link between colibactin and an increased CRC risk in human patients has been lacking[25]. Here, we present ClbB as independent factor associated with recent or prevalent dysplasia (aOR 7.97). The prevalence of ClbB in our study was similar between controls and UC patients, as opposed to a previous report showing significantly higher colibactin levels in IBD patients compared to controls.[26] However, most of those patients had active IBD, while all patients in our study were in endoscopic remission. The high prevalence of ClbB in control patients, but also other populations like FAP[9], suggests ClbB may be relevant for dysplasia development in multiple populations and warrants further research.

FadA and *Fusobacterium* presence were associated with an absence of (prior) dysplasia, in contrast to literature indicating a relation between CRC and increased FadA presence.[27],[28] FadA can bind to E-cadherin on epithelial cells and can promote inflammation and oncogenic processes.[27,29] The contrast between our results and the vast evidence of increased abundance of *Fusobacterium* and FadA in CRC underlines the hypothesis that this bacterium is associated mostly to later stages possibly by being attracted to tumor metabolites.[30] Moreover, FadA containing CRC isolates have failed to induce tumor formation in predisposed mice, while *E. coli pks+* has been shown to induce CRC through colibactin.[31] Another study showed that FadA could drive CRC in tumor prone mice but proposed that FadA only affects cancerous cells.[29] Moreover, *Fusobacteria* have variations in presence and expression of FadA[32], resulting in strain-rather than species-dependent oncogenic potential.[33] Recent literature even suggests that *Fusobacteria* supernatants induce expression of immunomodulatory *TGF*β*1 in vitro*, possibly by its production of butyrate.[34] Our results support that *Fusobacteria* do not predict early dysplasia and we even observed an inverse relationship of *Fusobacteria* and FadA in early carcinogenesis in UC.

Two other toxin encoding genes, Eae and BFT were not associated with dysplasia in UC patients. Eae, which has been shown to induce loss of DNA mismatch repair genes[35-37], did not occur without ClbB, possibly because both genes are found in *E. coli*. The effect of Eae was not significant and therefore is likely to play a minor role in dysplasia risk as opposed to ClbB. Previously, *B. fragilis* and BFT were reported to be over-expressed in tissue biopsies of UC patients with active disease and to be frequently present in the mucosa of patients with CRC and control patients.[8,19,38] Similar to *Fusobacterium, B. fragilis* without BFT has immunosuppressive effects through outer membrane vesicles and polysaccharide A. Hence, bacteria of the same species, but with different oncotrait expression, may have opposing effects on colonic tissue.[33]

Colonic biofilms have been associated with gastro-intestinal symptoms in patients with UC and other IBDs[7,8], as well as sporadic CRC and FAP. A study from Baumgartner et al. detected visible biofilms in 34% and microscopic biofilms in 79% in UC patients comparable to our microscopic biofilm rate of 72.2% in high-risk UC patients at study colonoscopy. These authors found a low biofilm prevalence of 6% in controls compared to the higher prevalence in our study (50%). Importantly, this might be explained by the different selection of controls in our study, also allowing patients with symptoms to be included. Moreover, biofilms were numerically more frequent and thicker in the right-sided colon compared to the more distal colon, were frequently persistent over time, and showed an association with epithelial hypertrophia in UC patients. Samples with biofilms were associated with a higher bacterial abundance, but a lower Shannon diversity than samples without biofilms[16], indicating the outgrowth of specific bacteria; *Clostridiales, Bacteroidales* and *Veillonellales* (adj-p=0.012) in metagenomes of controls and *Selenomonadales* and *Synergistales* in metagenomes of UC patients (adj-p=0.011). In a mouse model, biofilms from both CRC and control patients were carcinogenic,[39] however, our longitudinal data neither showed a significant association between biofilm presence and dysplasia risk in UC patients, nor showed that biofilms associated with high dysplasia risk are different from biofilms in low risk patients. We speculate that this is the consequence of biofilms harboring a multitude of bacterial species, not all associated with pro-inflammatory or oncogenic characteristics.

### Strengths and limitations

This study has multiple strengths. We extensively assessed biofilm presence over time as well as cross-sectionally in a prospective fashion, providing a link with clinical and histological presence of inflammation and dysplasia. Furthermore, our colonoscopy controls were carefully selected and had no abnormal findings during colonoscopy such as dysplasia and inflammation. In addition, this is the first study that assessed oncotrait presence in UC in relation with dysplasia. There are also limitations. Although we prospectively collected data from our patients, the additional longitudinal analysis of biopsies that were collected prior to the study colonoscopy has a retrospective nature and involved formalin fixed rather than methacarn fixed tissues which may affect sensitivity of biofilm detection. Finally, from our controls we do not have a longitudinal follow-up, and we are unable to predict if the presence of oncotraits in controls increases the risk for dysplasia.

## Conclusion

In conclusion, our results suggest that oncotrait presence within bacterial species may be crucial to understanding which patients are at risk for dysplasia development. This points towards a role for specific microbial functions rather than biofilms in general as potential risk factor for CRC. ClbB presence and FadA absence are were significantly and independently associated with dysplasia risk in UC patients and are therefore useful biomarkers. Since these markers are detectable in feces, they could be used to predict low-risk patients and thereby reduce the number of invasive endoscopic procedures for CRC prevention. More research is needed to see whether more oncotraits could be incorporated into fecal screening and whether such screening could aid beneficial effects of fecal transfers. Indeed, incorporation of ClbB fecal testing in a CRC detection model for the general population has shown promise and resulted in an improved sensitivity.[40,41] This emphasizes the potential for ClbB in future CRC risk stratification in UC and improving the discriminatory value for use in clinical practice.

## Data Availability

Raw sequencing data with human reads will not be publicly available because of General Data Protection Regulation (GDPR). Processed sequencing data are available upon request. Anonymized patient and research data that support the findings of this study are openly available in supplementary data.

## Abbreviations used in this paper

IBD: inflammatory bowel disease
UC: ulcerative colitis
PSC: primary sclerosing cholangitis
CRC: colorectal cancer
FDR: First degree relative
HGD: high-grade dysplasia
LGD: low-grade dysplasia
SSCAI: Simple Clinical Colitis Activity Index
OR: odds ratio
aOR: adjusted odds ratio
BFT: Bacteroides fragilis toxin
qPCR: quantitative PCR
n: number
IQR: interquartile range
IHC: Immunohistochemistry
FISH: Fluorescent *in situ*hybridization
SD: standard deviation
CI: Confidence interval

