## Supplementary Data 1 for "BACTERIAL ONCOTRAITS BUT NOT BIOFILMS ARE ASSOCIATED WITH DYSPLASIA IN ULCERATIVE COLITIS"

The potential of bacterial oncotraits and longitudinal biofilm presence to predict dysplasia in ulcerative colitis

*Carlijn Bruggeling^1^, Maarten te Groen^2,^*, Daniel Garza^3,^*, Famke van Heeckeren tot Overlaer^1^, Joyce Krekels^1^, Basma-Chick Sulaiman^1^, Davy Karel^1^, Athreyu Rulof^1^, Anne R. Schaaphok^1^, Daniel Hornikx^1^, Iris D. Nagtegaal^1^, Bas E. Dutilh^4,5^, Frank Hoentjen^3,6^, Annemarie Boleij^1^*

Supplementary Methods

### Fluorescent in situ hybridization (FISH)

*Tissue processing*

Methacarn fixed left- and right-sided biopsies in paraffin embedded blocks were sliced into 30 sections of 4 µm, of which the 10^th^, 20^th^ and 30^th^ were analyzed with FISH. Another round of 30 sections was performed if no biofilms were observed. In addition, also formalin fixed paraffin embedded material collected for standard diagnostic procedures was collected and processed for immunostainings and FISH(retro- and prospectively).

*FISH*

Sections were deparaffinized through 5-minute incubations in xylene (2 times) and rehydrated in alcohol (2 times). Sections were air-dried before adding the probe (3 ng/µL) in hybridization buffer (0.9 M NaCl, 20 mM Tris-HCl, pH 8.0, 0.01% sodium dodecyl sulfate). A universal bacterial probe EUB338 was used (5’-Cyanine3- GCTGCCTCCCGTAGGAGT-Cyanine3-3’) and non-EUB338 (5’-Cyanine3- ACTCCTACGGGAGGCAGC-3’) was used as negative control [ref]. Sections were incubated overnight in a humidified chamber at 46°C. Subsequently, sections were washed 3 times for 5 minutes at 48°C in washing buffer (0.9M NaCl, 20 mM Tris-Cl, pH 8.0), followed by one minute in ice-cold water and one minute in PBS. Sections were mounted with Prolong Gold Antifade medium with DAPI (Thermo Fisher Scientific, P36931). Slides were scored on a fluorescence light microscope (Leica DM4000).

### Immunohistochemistry

Sections were stained using standardized procedures of the diagnostics division of Pathology in the Radboudumc. Four µM thick sections were deparaffinized in xylene and rehydrated in graded series of alcohol. Sections were incubated at 96 °C for 10 minutes in EDTA buffer (pH 9.0) in the Lab Vision PT-module). Incubation with 3% H2O2/methanol solution was performed in the automated Lab Vision immunostainer (Immunologic), after which sections were incubated for 1 hour at room temperature with anti-Ki67 antibody (Mib-1, 1:50, DAKO) in normal antibody diluents (Immunologic). BrightVision (Immunologic) with bright- 3,3′-diaminobenzidine (DAB) (Immunologic) were used for visualization, after which sections were counterstained with hemaoxylin and mounted in an automated slide-stainer and coverslipper (Tissue-tek prisma). Tonsil, gut, liver and pancreas were used as a control for each run. All slides were scanned using the Panoramic scanner and Viewer (3DHISTECH Ltd.).

### Thickness of the mucuslayer

Sections were stained with a Periodic acid-Schiff (PAS) and scanned and scored in the Panoramic Scanner and viewer (3DHISTECH Ltd.). Maximum, median and minimal mucus thickness was scored based on 10 measurements every 200 µm of the epithelium.

### Multiplex quantitative PCR for oncotraits

A multiplex PCR detection of *Bacteroides fragilis* toxin (BFT), Colibactin (ClbB gene), Intimin (Eae)*,* and *Fusobacterium* adhesin A (FadA) was performed. Taqpath multiplex 1 Step Mastermix was supplemented with 45nM forward and reverse primer and 12.5 nM Probe for detection of BFT, Intimin, FadA and ClbB. Plates were run at 2 minutes 50°C, 10 minutes 95 °C, followed by 40 cycles of 15 sec 95 °C, 1 min 60 °C (Applied biosystems 7500 fast system). An equal mix of probes for BFT, Eae, FadA and ClbB was used for generation of a standard curve (2.5 ng/µL to 0.2 ng/µL). Negative controls were a mixture of equal amounts of DNA from representative strains without BFT, Intimin, FadA and Colibactin at 2.5 ng/ul. Each fecal DNA sample was run in duplicate at 25 and 2.5 ng of DNA input. A signal was considered positive when the fluorescence signal exceeded two times the magnitude of the standard deviation of the baseline signal^1^.

| Gene | Forward primer | Reverse primer | Probe | Label |
| --- | --- | --- | --- | --- |
| BFT | GGATAAGCGTACTAAAATACAGCTGGAT | CTGCGAACTCATCTCCCAGTATAAA | CAGACGGACATTCTC | 5’JOE,  3’OQA |
| Intimin (Eae) | ATCTCTGAACGGCGATTACG | TCCAGTGAACTACCGTCAAAG | ACCAGGCTTCGTCACAGTTGCAG | 5’ROX,  3’OQC |
| FadA | AATTTGGGAGGTAACAAAAATGAAAAA | GCTAAGTTTTGGTATTCAGCATCTAG | ACCTACTAAACTTGCTGCATCAGTTGCT | 5’6FAM, 3’OQA |
| Colibactin (ClbB) | GGCGTAATCCCCAGATCAATC | GATGAGGATACGCGGGATATTG | TGTTCTGTGTCGAATACAGCCTAGCG | 5’TAMRA, 3’OQB |

### Metagenomics

Shotgun metagenomic sequencing was performed as reported before^2^. A total of 275 biopsies from UC and control patients were send to Novogene Bioinformatics Technology Co., Ltd in Hong Kong for sequencing using the low input NEBnext library preparation and paired-end sequencing with the Illumina Novaseq 6000 using 350 bp insert size and a read length of 150 bp. 1.2 GB output data in FastQ format was guaranteed per sample. Samples were measured for DNA concentration (Qubit), construct length and a quality check was performed on the library preparation. Ten samples were not sequenced due to failed library preparation resulting in data of 265 biopsies of which 122 right-sided biopsies, 119 left-sided biopsies, and 24 samples from neoplastic or inflamed sites (Supplementary Table 2).

### Bioinformatics analysis

Quality control, trimming, and removal of adaptors was performed using FastQC version 0.11.9 and trimmomatic version 0.35. An assembly dataset was generated by filtering out the human reads using BBMap version 38.84 with the GRCh38 version of the human genome. Filtered reads were assembled with metaSPAdes version 3.13.1. The taxonomic classification of contigs was determined with CAT v. 4.6^3^ using the NCBI NR as database for taxonomic assignments. bwa version 0.7.17 and samtools version 1.9 were used to map all the reads to the classified contigs and the human genome and to estimate the coverage statistics. Accuracy is estimated by predicting the presence of biofilm with a logistic regression using K-fold cross validation with one thousand independent iterations. In each iteration, a random subset of the data is used to fit a logistic regression model while another random subset is used to predict the label (e.g. biofilm presence, right-sided, or high-risk patients) with the fitted parameters. The final accuracy is computed as the average of predictions on each iteration. The process is then repeated with random labels to estimate an empirical p-values. This p-value is the proportion of times that the predictions with random labels are equal or more accurate as the predictions with the true labels. The positive and negative associations of each taxa level were defined with the Analysis of Composition of Microbiomes method (ANCOM)^4^ using permutative ANOVA as the statistical test with 1000 permutations and adjusting the p-values with the Benjamini-Hochberg false discovery rate (alpha<0.05 was considered significant).

1.	Pfaffl MW. A new mathematical model for relative quantification in real-time RT-PCR. Nucleic Acids Res 2001;29:e45-e45.

2.	Bruggeling CE, Garza DR, Achouiti S, et al. Optimized bacterial DNA isolation method for microbiome analysis of human tissues. Microbiologyopen 2021;10:e1191.

3.	von Meijenfeldt FAB, Arkhipova K, Cambuy DD, et al. Robust taxonomic classification of uncharted microbial sequences and bins with CAT and BAT. Genome Biol 2019;20:217.

4.	Mandal S, Van Treuren W, White RA, et al. Analysis of composition of microbiomes: a novel method for studying microbial composition. Microbial Ecology in Health and Disease 2015;26:27663.

Supplemental figures

### Supplemental figure 1: DAGs model for causal inference

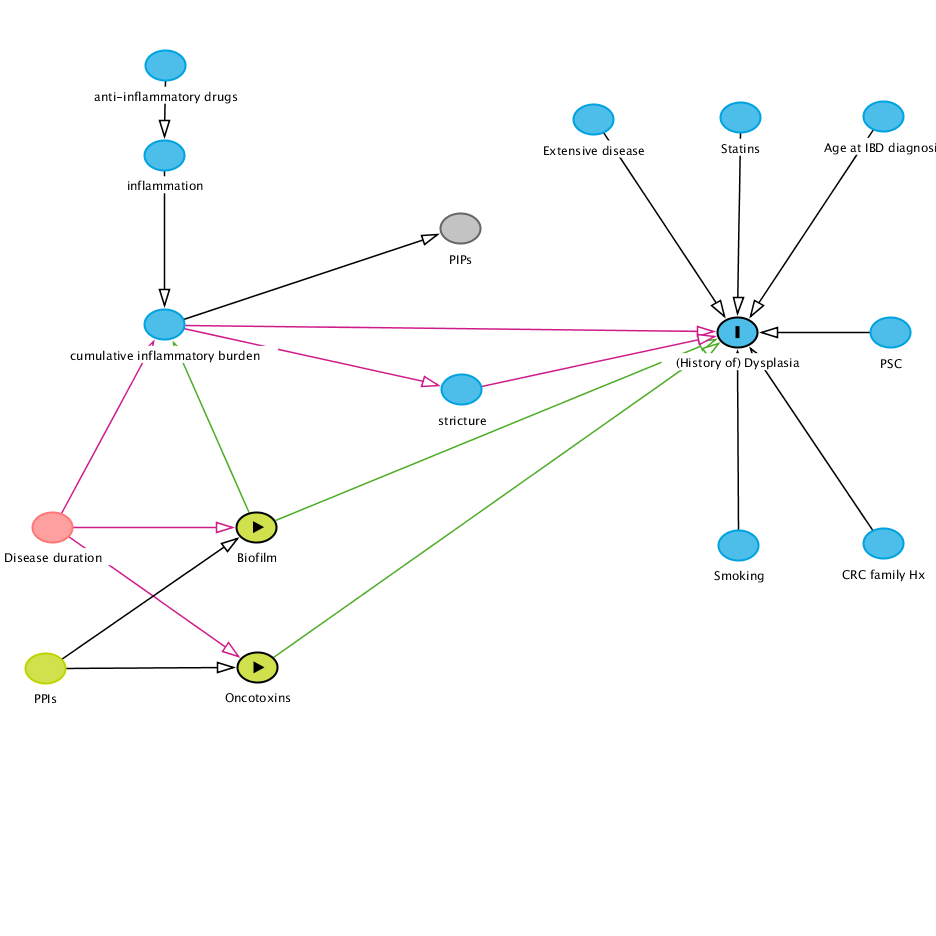

### Supplemental Figure 2: Biofilm area in UC-patients and controls

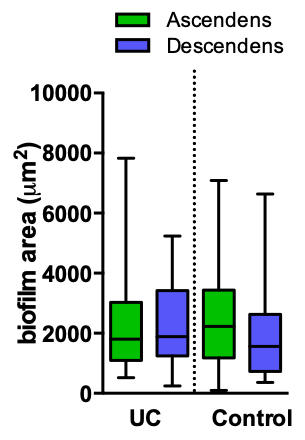

### Supplementary Figure 3: Correlation between mucus thickness and biofilm thickness

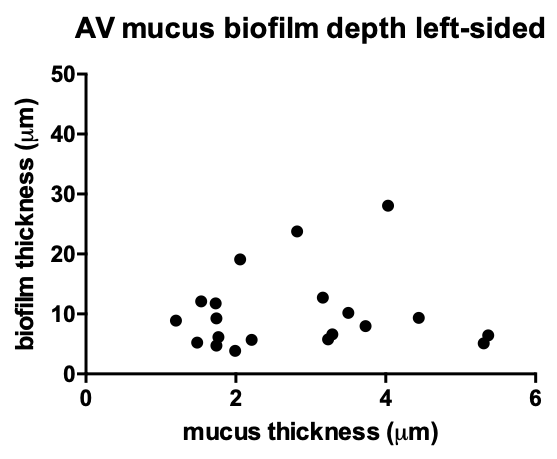

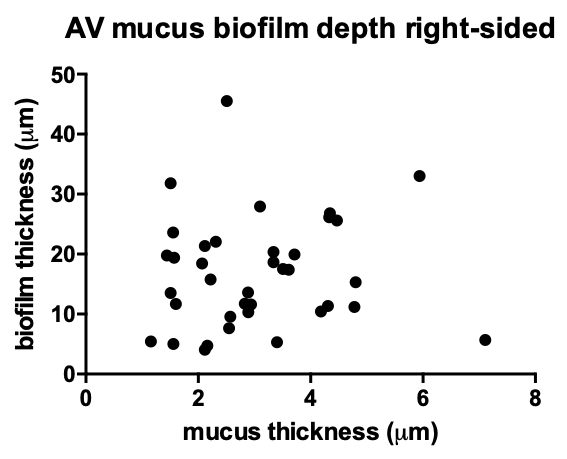

### Supplementary Figure 4: Longitudinal biofilms in patient with and without dysplasia

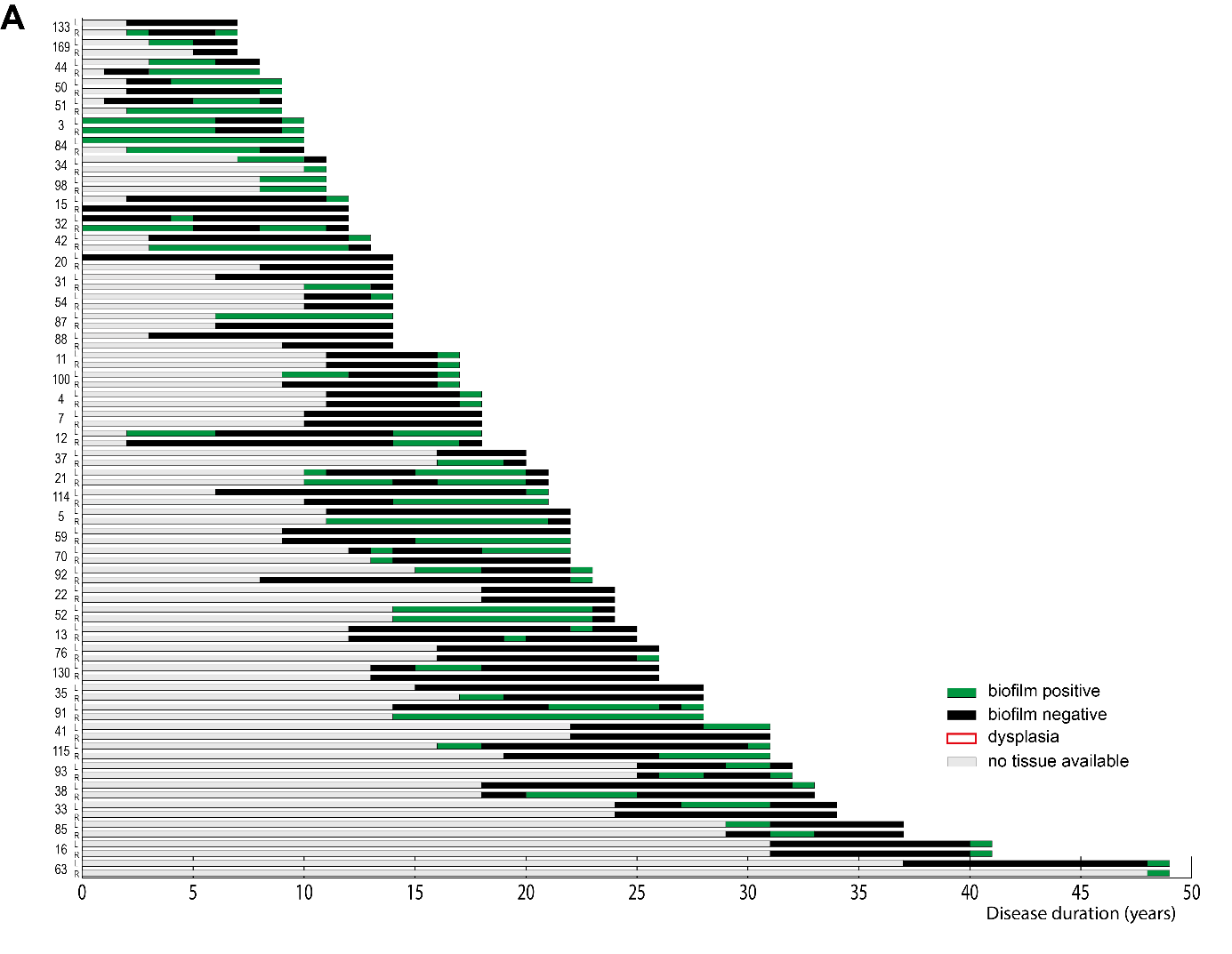

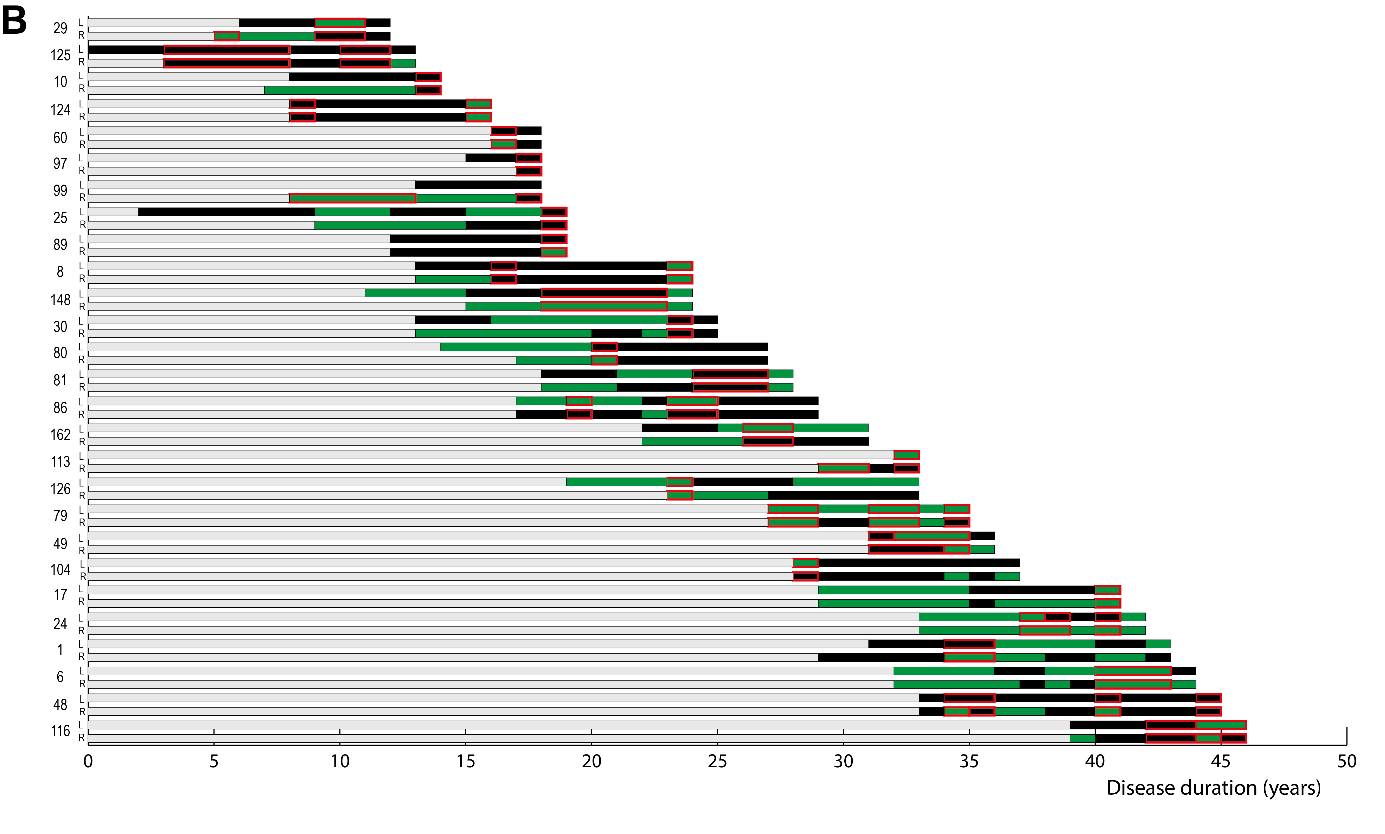

***Longitudinal biofilm presence per UC-patient.***

*A) UC-patients without dysplasia, and B) UC-patients with dysplasia. For each patient left (L) and right-sided (R) colonoscopies are displayed with disease duration on the x-axis. Data are sorted based on disease duration. Green bars represent biofilm positive episodes, black bars represent biofilm negative episodes, red squares indicate episodes with dysplasia present.*

### Supplementary Figure 5: Risk associations of metagenomes at order level with dysplasia

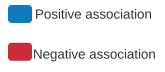

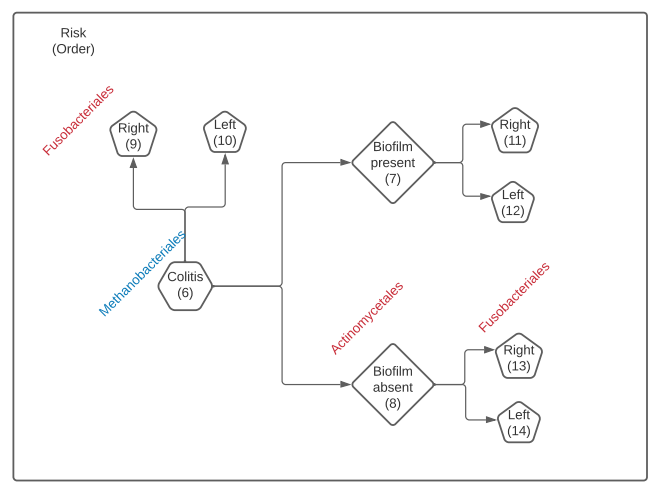
